# Identification of Plasma Growth Factors and Cytokines as Diagnostic Biomarkers for Lafora Disease

**DOI:** 10.1101/2025.04.29.25326186

**Authors:** Mireia Moreno-Estellés, María Machio, Laura González, Marta Albuixech, Laura Abraira, Manuel Quintana, Manuel Toledo, Marina P. Sánchez, José M. Serratosa, Pascual Sanz

## Abstract

Lafora progressive myoclonus epilepsy (LD, OMIM#254780, ORPHA:501) is an ultra-rare and severe autosomal recessive neurological disorder that typically manifests in early adolescence. It is characterized by the accumulation of insoluble forms of aberrant glycogen in the brain and peripheral tissues. Given the urgent need for reliable tools for diagnosis and to monitor disease progression, in this work, we aimed to identify reliable biomarkers in minimally invasive fluids, which could also provide valuable insights into the natural history of the disease. Plasma-EDTA samples from eleven LD patients and healthy controls were analyzed to identify potential biomarkers of LD. Eleven cytokines and growth factors were identified to be significantly reduced in LD patient samples compared to healthy controls. Among these, four mediators [platelet-derived growth factor subunit B (PDGF-BB), epidermal growth factor (EGF), brain derived growth factor (BDNF), and macrophage migration inhibitory factor (MIF)], exhibited the greatest fold change between the groups and were further validated. Receiver operating characteristic (ROC) analysis confirmed the robust diagnostic potential of these four proteins. Notably, the levels of these proteins were reduced even in asymptomatic patients, suggesting potential utility for early diagnosis. Given the minimally invasive nature of plasma sampling and the straightforward quantification via ELISA assays, these biomarkers hold strong promise for rapid translation to the clinic, potentially enhancing early diagnosis in LD patients. Our findings offer a promising step toward the development of accessible, non-invasive diagnostic tools for LD.,.

**Key points:**

- Several plasma biomarkers associated with Lafora disease (LD) have been identified.
- These biomarkers are reduced in LD patients compared to healthy controls.
- Reduced levels of these biomarkers were detectable even in asymptomatic patients.
- Their reduced levels may contribute to the neurological alterations observed in LD.

## INTRODUCTION

Lafora disease (LD, OMIM#254780, ORPHA:501) is an ultra-rare and fatal autosomal recessive form of progressive myoclonus epilepsy. LD typically manifests in early adolescence and is characterized by the accumulation of insoluble deposits of aberrant glycogen (polyglucosans) in the brain and peripheral tissues (Lafora and Glueck, 1911). More recently, neuroinflammation has been defined as a second hallmark of LD in mouse models (Lahuerta et al., 2020), (Rubio et al., 2023). LD results from mutations in the *EPM2A* gene, which encodes the glucan phosphatase laforin, or in the *EPM2B* gene, which encodes the E3-ubiquitin ligase malin. Laforin and malin form a functional complex where laforin recognizes substrates to be subsequently ubiquitinated by malin (Solaz-Fuster et al., 2008). This shared molecular mechanism likely explains the similar clinical presentations observed in patients carrying mutations in either gene. The laforin-malin complex negatively regulates glycogen synthesis by modulating the activity of glycogen synthase (Vilchez et al., 2007), thereby providing a mechanistic basis for the enhanced glycogen synthesis observed in LD patients.

Clinically LD is characterized by generalized tonic-clonic seizures, myoclonus, absences, and visual hallucinations. The disease progresses rapidly with patients experiencing cognitive deterioration, dementia, and increased seizure frequency, often leading to death within 5 to 15 years after onset due to complications such as status epilepticus or aspiration pneumonia (Pondrelli et al., 2021). The rarity of the disease (prevalence of less than 4/1,000,000 individuals) has hindered comprehensive studies of its natural history. However, a recent analysis of 298 LD cases provided valuable insights into key clinical milestones, including the mean age of onset (approximately 13 years), median survival time (around 11 years), and median time to loss of autonomy time (approximately 6 years), among other parameters (Pondrelli et al., 2021). These findings complement earlier prospective studies using electroencephalographic (EEG) biomarkers and Clinical Disability Progressive Scale (CDPS), which delineate five clinical stages of LD: stage 0, asymptomatic; stage I, with visual seizures only; stage II, with mild cognitive decline; stage III, with an established dementia and status epilepticus; stage IV, with myoclonic encephalopathy; and stage V, with progressive neurological deterioration, eventually leading to death from respiratory failure (Delgado-Escueta et al., 2015; Cure Lafora Epilepsy Meeting, Istanbul, Turkey, https://chelseashope.org/wp-content/uploads/2018/08/Istanbul-Meeting-Report-Escueta.pdf) (Kim et al., 2021).

To monitor disease progression, there is an urgent need to identify surrogate biomarkers associated with LD. These biomarkers, whether diagnostic or prognostic, could predict disease manifestations, track progression, and assess the efficacy of emerging therapies. To date, to our knowledge, only imaging techniques such as positron emission tomography (PET), magnetic resonance spectroscopy and imaging (MRI), and proton magnetic resonance spectroscopy (MRS) have been used to monitor the progression of LD patients (Burgos et al., 2020), (Chan et al., 2024), (d’Orsi et al., 2023), (Muccioli et al., 2020),

Using animal models of LD, alternative potential biomarkers have been explored. For example, metabolomics analyses have revealed differences in brain metabolites between LD and control mice (Brewer et al., 2019), (Markussen et al., 2024). Additionally, our group recently identified elevated levels of specific inflammatory cytokines in the serum samples of LD mice compared to controls (Rubio et al., 2023).

The aim is this work is to identify in the plasma of LD patients some determinants that could serve as diagnostic biomarkers of LD.

## MATERIALS AND METHODS

### Study design

Plasma-EDTA samples from eleven LD patients and thirteen healthy subjects were provided by Fundación Jiménez Díaz, Hospital Vall d’Hebron, and the CIBERER Biobank. Healthy control samples were collected in two different recruitments (H samples). Samples from five other monogenic epilepsies (Dravet and GLUT1 deficiency, OME samples) were included to assess the specificity of the newly identified biomarkers **(Table 1)**. Samples were processed following standard operating procedures and with the appropriate approvals from the corresponding Ethics and Scientific Committees (see Ethics approval statement). Healthy and OME subjects were selected to match the age range of the LD patients. When possible, LD samples were collected from the same patients during three consecutive visits to the Neurology Department, at different intervals (T1 to T3). Male and female LD patients with mutations in either the *EPM2A* or *EPM2B* genes were included in the study **(Table 1)**. For each LD patient, a comprehensive battery of cognitive and behavioral assessments (including seizures, dementia, and gait impairment) was conducted **(Supplementary Table S1**), as well as the identification of the specific LD mutation. The Clinical Disability Progressive Scale (CDPS) (Delgado-Escueta et al., 2015; Cure Lafora Epilepsy Meeting, Istanbul, Turkey, https://chelseashope.org/wp-content/uploads/2018/08/Istanbul-Meeting-Report-Escueta.pdf) (Kim et al., 2021), was used to determine the stage of the disease at each visit **(Supplementary Table S1**). Data on ongoing treatments were also recorded.

**Table 1.** Age range and gender of the subjects analyzed in this work. F: female; M: male. T: time of the visit. $, healthy samples used in the Proteome Profiler Human Cytokine Array kit; LD, Lafora disease; OME, other monogenic epilepsy.

| Healthy subjects | Age range (years)/ gender | Patient and (mutation) | Age range (years) at the beginning of the study (T1)/gender | Samples obtained during the study |
| --- | --- | --- | --- | --- |
| H1<sup>\$</sup> | 26-30/ F | LD1 (EPM2A) | 16-20 / F | T1, T2, T3 |
| H2<sup>\$</sup> | 31-35/ M | LD2 (EPM2A) | 16-20 / M | T1 |
| H3<sup>\$</sup> | 21-25 / F | LD3 (EPM2B) | 16-20 / F | T1, T2, T3 |
| H4<sup>\$</sup> | 31-35/ M | LD4 (EPM2A) | 16-20 / M | T1, T2 |
| H5<sup>\$</sup> | 21-25 / F | LD5 (EPM2A) | 31-35 / M | T1, T2 |
| H6<sup>\$</sup> | 26-30 / M | LD6 (EPM2A) | 6-10 / F | T1, T2, T3 |
| H7<sup>\$</sup> | 21-25 / F | LD7 (EPM2A) | 11-15 / M | T1, T2, T3 |
| H8<sup>\$</sup> | 21-25 / M | LD8 (EPM2B) | 16-20 / F | T1 |
| H9 | 21-25 / F | LD9 (EPM2A) | 11-15 / M | T1 |
| H10 | 26-30 / F | LD10 (EPM2B) | 11-15 / F | T1, T2 |
| H11 | 26-30 / M | LD11 (EPM2B) | 11-15 / F | T1, T2 |
| H12 | 21-25 / F |  |  |  |
| H13 | 26-30 / M |  |  |  |
|  |  | OME01 (SCNA1) | 16-20 / M | T1 |
|  |  | OME02 (SCNA1) | 16-20 / M | T1 |
|  |  | OME03 (SCNA1) | 26-30 / M | T1 |
|  |  | OME04 (SLC2A1) | 26-30 / M | T1 |
|  |  | OME05 (SLC2A1) | 26-30 / M | T1 |

Blood samples were collected in EDTA tubes, centrifuged at 800xg for 10 min at room temperature, and plasma aliquots were frozen at -80ºC until use.

### Biomarker validation

Selected mediators were validated using specific ELISA assays, including PDGF-BB (Abcam, ab184860), EGF (Abcam, ab217772), BDNF (Cloud-Clone Corporation, SEA011Mi), MIF (R&D systems, DMF00B), CXCL10/IP10 (Abcam, ab173194), S100B (Abcam, ab234573), and CCL20/MIP3a (R&D systems, DM3A00). Manufacturer protocols were followed in each case.

### High throughput screening of inflammatory mediators

Plasma-EDTA samples from LD patients and healthy controls were analyzed using the Proteome Profiler Human Cytokine Array kit (R&D systems). This array consists of 111 different captured antibodies spotted on a nitrocellulose membrane for the detection of multiple cytokines, chemokines, growth factors, and other soluble proteins. The intensity of the signals was analyzed using the *Quick Spot* software provided by the manufacturer (**Supplementary Table S2**). The analysis of the differential expression between healthy and LD samples was performed by using the *FLASKI* software (http://flaski.age.mpg.de). In this work, we focus our attention on mediators with a fold change between healthy and LD samples greater than 2 or less than 0.5 for further analysis.

### Protein interactions

Putative interactions among the selected mediators were assessed using *STRING v12*.*0* software (https://string-db.org/), which integrates data from numerous sources, including experimental studies, computational predictions, and publicly available literature.

### Statistical analysis

Statistical differences between the groups were analyzed using unpaired, non-parametric Mann-Whitney t-tests, using GraphPad Prism version 6.0 statistical software (La Jolla, CA, USA). Results are expressed as medians with ranges. Statistical significance was defined as P-values ****p<0.0001, ***p<0.001 and *p<0.05. Receiver operating characteristic (ROC) analysis was performed for the validation of the four putative biomarkers to assess their capability to diagnose LD by plotting sensitivity versus (1-specificity), using the IBM SPSS statistical package (version 25; SPSS Inc., Armonk, NY, USA). The area under the ROC curve (AUC) was calculated and Youden Index was used to determine optimal cut-off points for each biomarker.

## RESULTS

### 1. Analysis of CXCL10, S100B and CCL20 protein levels

In a recent study, we reported elevated levels of CXCL10, S100B, and CCL20 proteins in the blood serum of a mouse model of LD (*Epm2b-/-* mice) aged 12 months or older (Rubio et al., 2023). To assess whether these proteins were also elevated in human samples from LD patients, we measured their levels using ELISA assays in plasma samples from seven LD patients (LD1 to LD7) and four age-matched healthy controls (H1 to H4) (**Table 1 and Supplementary Table S1**). In the case of CXCL10, we observed higher levels of this chemokine in the LD samples compared to controls (median 64 pg/ml in healthy vs 70 pg/ml in LD patients), but this difference was not statistically significant (p=0.494; **Supplementary Fig. S1A)**. No significant differences were found either in the levels of S100B (median 1 ng/ml for healthy vs 1 ng/ml for LD patients; p=0.999; **Supplementary Fig. S1B)** or in the case of CCL20 proteins (median 31 pg/ml for healthy vs 1 pg/ml for LD patients; p=0.530; **Supplementary Fig. S1C)**. These results indicate that the levels of CXCL10, S100B, and CCL20 are not reliable biomarkers for LD in humans.

### 2. Analysis of human plasma samples using high throughput arrays

To expand our search for potential biomarkers of LD, we analyze the cytokine profile of plasma samples from LD patients using the Proteome Profiler Human Cytokine Array kit (see Methods). This analysis included eight healthy samples (marked with ^$^ in **Table 1**; 4 men and 4 women) and the first extraction (T1) from eleven LD samples (4 men and 7 women), as described in **Table 1**.

The intensity of the signals was analyzed using the *Quick Spot software* provided by the manufacturer. In **Supplementary Table S2** we show the average intensity of all the spots, the fold change value between healthy and LD samples, and the corresponding p-values. In **Fig. 1A** we show the heatmap corresponding only to those proteins with a fold change value between healthy and LD samples greater than 2 or lower than 0.5. A clear distinction between healthy and LD samples was observed for these proteins. In the right panel of **Fig. 1A**, we indicate the fold change of the average intensity of each protein and the corresponding p-values. In this approach, we also confirmed the absence of significant differences in the levels of CXCL10 and CCL20 proteins between healthy and LD samples (see values in grey in **Supplementary Table S2**; the human array did not contain S100B antibodies).

**Fig. 1.**
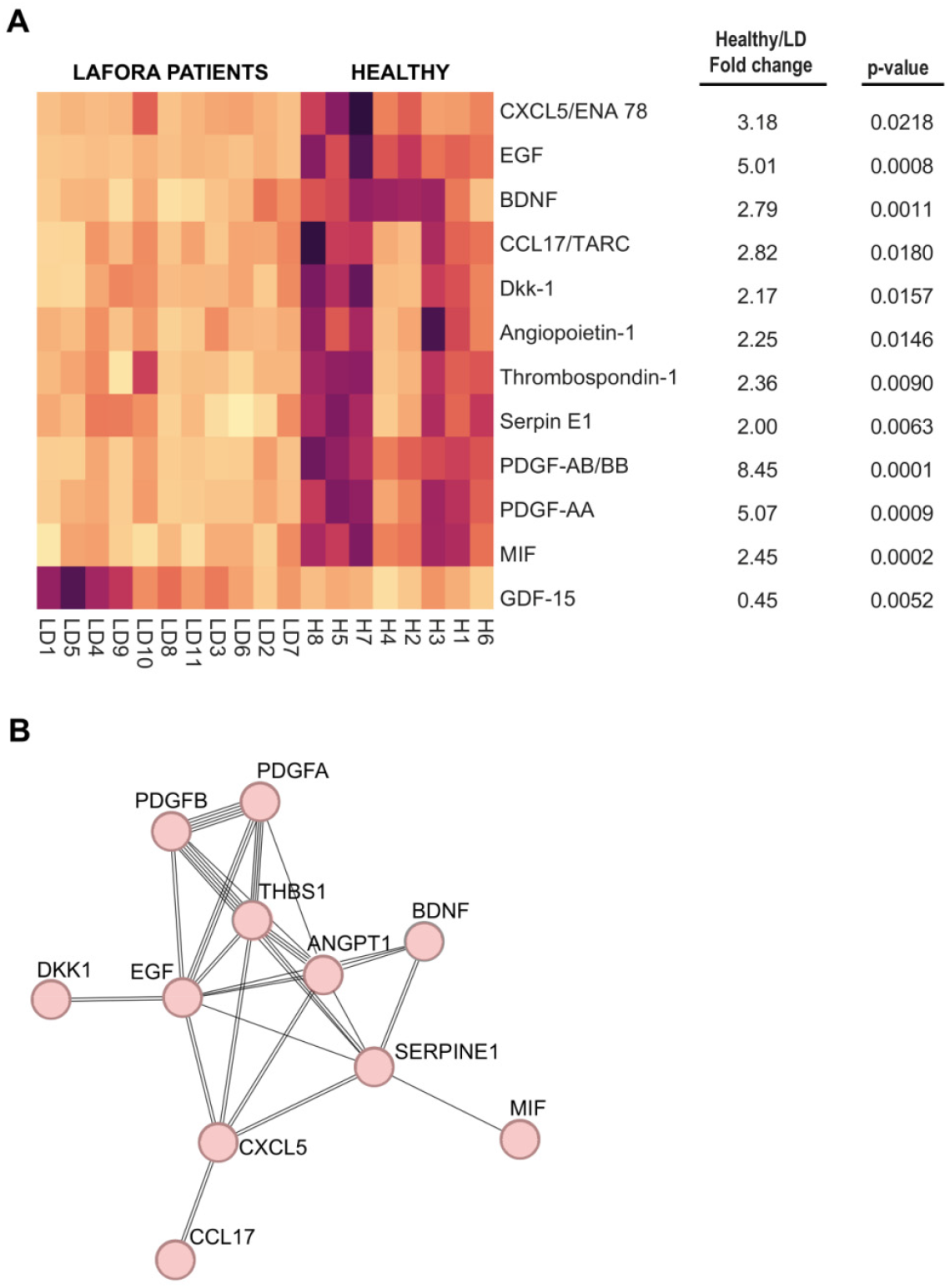
Differentially expressed proteins in Lafora disease patients and protein-protein interaction analysis. **A)** Heatmap showing the levels of different proteins detected using the Proteome Profiler Human Cytokine Array kit (R&D systems, see Methods). Plasma samples from eight healthy subjects and eleven LD patients were analyzed according to the manufacturer’s instructions. The intensity of the signals was analyzed using the Quick Spot software provided by the manufacturer. Heatmap includes only proteins with a fold change value between healthy and LD samples greater than 2 or lower than 0.5, along with their corresponding p-values. **B)** STRING analysis of the proteins identified in Fig. 1A. Lines indicate possible protein-protein interactions, with a higher number of lines representing stronger predicted interactions.

In the LD samples, we only observed a significant increase in the case of GDF-15, a protein belonging to the transforming growth factor beta superfamily, which plays a role in regulating inflammatory pathways and is upregulated in cardiovascular and neuroplastic disorders (Zimmers et al., 2005), (Wollert et al., 2007), (Rochette et al., 2020) (**Fig. 1A and Supplementary Table S2**). However, it has been reported that treatment with metformin increases the levels of GDF-15 (Coll et al., 2020), and since the LD patients with higher levels of GDF-15 in the array (e.g., LD1, LD4, LD5, LD08, LD9, LD10, and LD11; **Fig. 1A**) were undergoing metformin treatment (see **Supplementary Table S1**), we decided to defer GDF-15 for further analysis.

Interestingly, we observed significant decreases in the levels of growth factors and other mediators in the LD samples **(Fig. 1A and Supplementary Table S2)**. Among them, we found particularly striking the decrease in the levels of platelet-derived growth factor AB/BB (PDGF-AB/BB; 8.45 fold decrease), PDGF-AA (5.07 fold decrease), and epidermal growth factor (EGF; 5.01 fold decrease). In addition, we found a decrease in the levels of chemokine CXCL15/ENA78 (3.18 fold decrease); chemokine CCL17/TARC (2.82 fold decrease); brain-derived neurotrophic factor (BDNF; 2.79 fold decrease); macrophage migration inhibitory factor (MIF, 2.45 fold decrease); thrombospondin-1, a glycoprotein that mediates cell-to-cell and cell-to matrix interactions (2.36 fold decrease); angiopoietin-1, a glycoprotein that mediates interactions between the endothelium and surrounding matrix (2.25 fold decrease); Dkk-1, an antagonist of the Wnt/b-catenin pathway (2.17 fold decrease); and endothelial plasminogen activator inhibitor 1 (serpin E-1), which is an inhibitor of tissue-type plasminogen activator (tPA) and urokinase (uPA), and it is regulator of cell migration (2.0 fold decrease) (the description of the proteins has been made according to Uniprot: https://www.uniprot.org/). Then, we used the STRING v12.0 software to analyze the possible functional interaction between these 11 mediators. This analysis pointed out a potential hub of interactions involving PDGF-AA, PDGF-BB, EGF, and thrombospondin-1 (**Fig. 1B**). Notably, the levels of none of these markers were decreased in samples from *Epm2b-/-* mice (Rubio et al., 2023), pointing out again to differences between LD mice and LD patients.

Among all these proteins, we decided to focus our attention on those with the highest average fold change in healthy vs LD samples and lowest p-values, selecting PDGF-AB/BB, EGF, BDNF, and MIF for further validation **(Fig. 1A and Supplementary Table S2)**.

### 3. Confirmation of the levels of PDGF-AB/BB, EGF, BDNF, and MIF by ELISA assays

Next, we validated the levels of the four selected proteins mentioned above using individual ELISA assays. We analyzed eleven samples from both the healthy donors and the LD patients. In the case of the LD patients, since in some of them we obtained several samples corresponding to different times of the progression of the disease (T1 to T3), we plotted in **Fig. 2** the values corresponding to the first time of extraction (T1) (see **Supplementary Table S3** for the complete dataset of values).

**Fig. 2.**
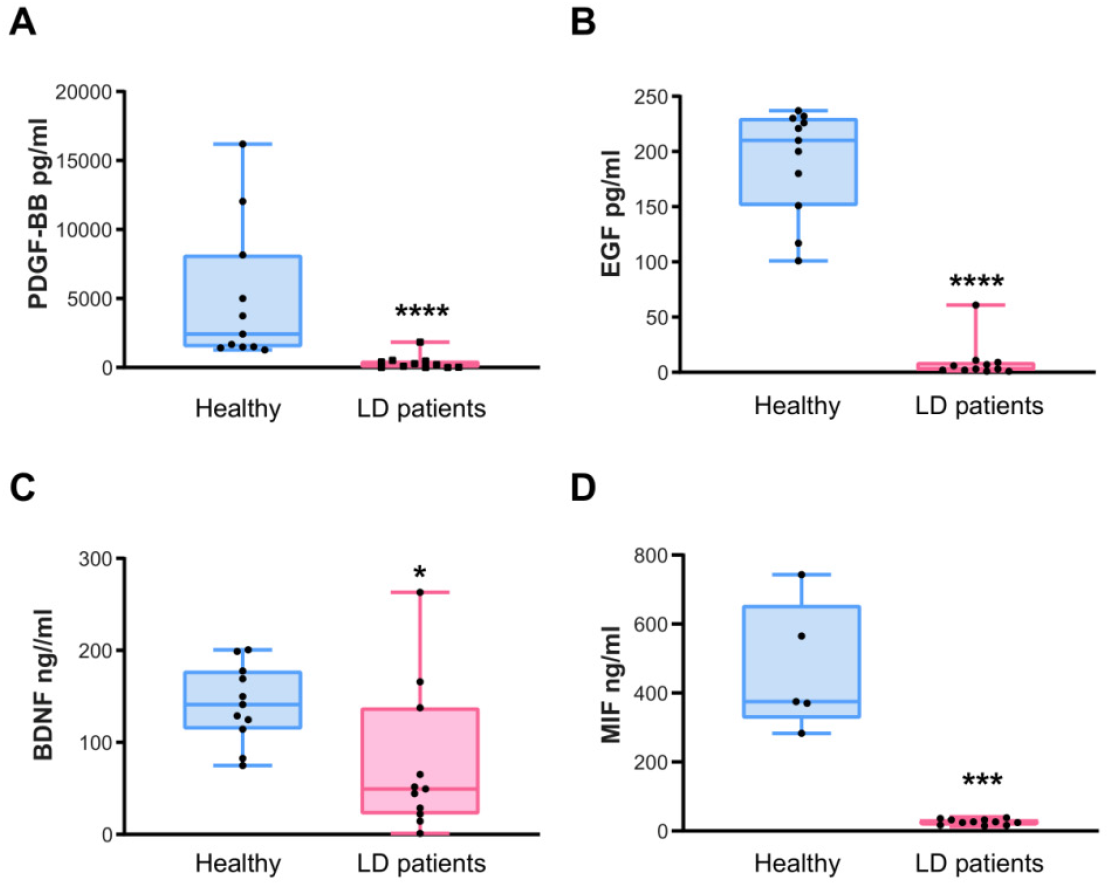
ELISA validation of the most significant proteins differentially expressed in LD samples. Plasma levels of PDGF-BB (A), EGF (B), BNDF (C), and MIF (D) in samples from eleven healthy subjects (pale blue) and eleven LD patients (pale pink) were analysed by ELISA assays (for the MIF analysis, only five healthy subjects were included). Results are expressed as a median with a range. Statistical differences between the groups were assessed using the Mann-Whitney non-parametric t-test. P-values have been considered as ****p<0.0001, ***p<0.001 and *p<0.05.

First, we confirmed significantly lower levels of PDGF-BB protein in all the samples of LD patients in comparison to healthy controls **(Fig. 2A)** (median of 2,424 pg/ml in healthy samples vs. 215 pg/ml in LD patients; p-value<0.0001). Secondly, we verified the reduction in the levels of EGF protein in all the samples of LD patients **(Fig. 2B)** (median of 210 pg/ml in healthy samples vs 3 pg/ml in LD patients; p-value<0.0001). In the case of BDNF, the decrease in the levels of this protein was also statistically significant in LD samples **(Fig. 2C)** (median of 141 ng/ml in healthy samples vs 49 ng/ml in LD patients; p-value=0.0128). Finally, in the case of MIF protein, we also found a decrease in the levels of this protein in LD samples **(Fig. 2D)**, which was statistically significant (median of 375 ng/ml in healthy samples vs 26 ng/ml in LD samples; p-value=0.0005).

### 4. Correlation of PDGF-BB, EGF, BDNF, and MIF levels with the clinical presentation of LD patients

To determine whether these four proteins could be considered biomarkers of LD, we performed a receiver operating characteristic (ROC) analysis to assess their predictive accuracy. ROC curves analysis shown in **Fig. 3** showed excellent diagnostic capabilities for PDGF-BB, EGF, and MIF, with elevated values of AUC in all the cases (95.9% for PDGF-BB, 100% for EGF, and 100% for MIF). The optimal cut-off values were <904 pg/ml for PDGF-BB, <81 pg/ml for EGF, and <160 ng/ml for MIF. BDNF exhibited a moderate predictive value (AUC was only 81% and the optimal cut-off value was <70 ng/ml).

**Fig. 3.**
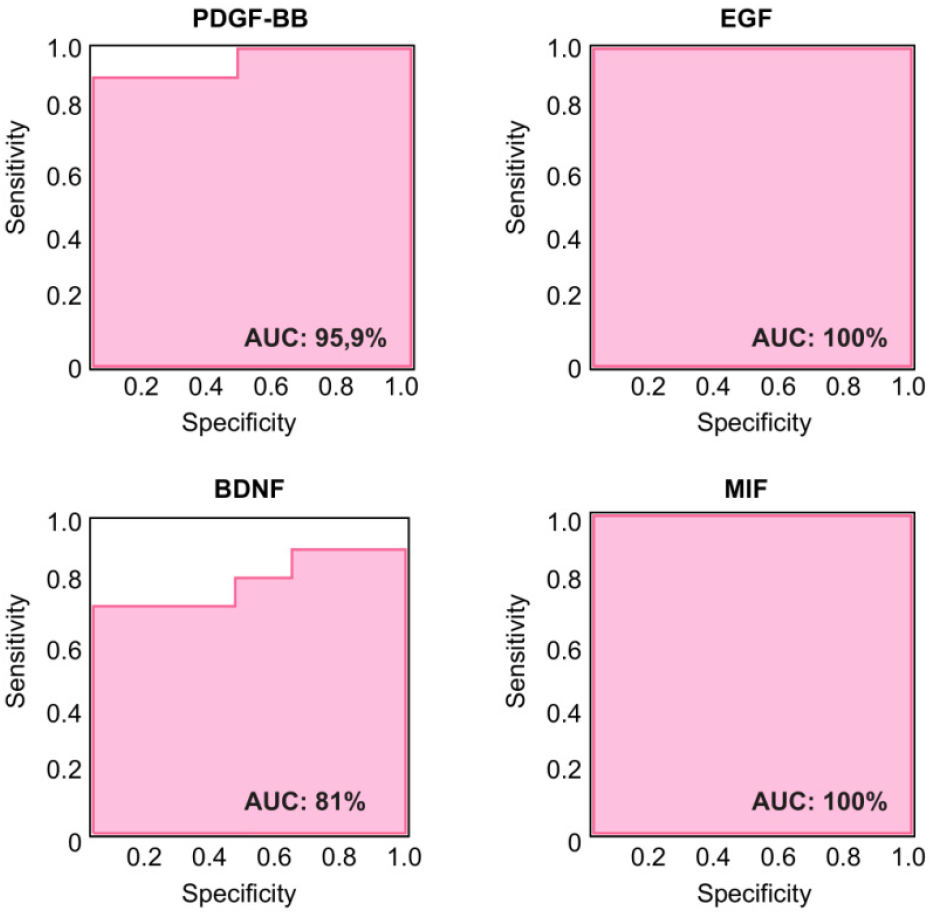
Receiver operating characterization (ROC) analysis of PDGF-BB, EGF, BDNF, and MIF. The ROC curves corresponding to the analysed proteins were generated to predict their potential use as biomarkers. The area under the curve (AUC) is indicated in each case.

Next, we studied then the possible correlation between the levels of these biomarkers and the progression of the disease in each LD patient. To this end, we plotted the absolute values of all the extractions of the LD samples **(Fig. 4)**. In the case of PDGF-BB, although the levels found in healthy samples showed substantial variability, the levels found in LD samples showed reduced values in most of the cases. In the case of EGF and MIF proteins, the levels found in all LD samples were always reduced in comparison with those of the healthy subjects. However, in the case of BDNF, some values of LD samples were as high as some found in healthy controls.

**Fig. 4.**
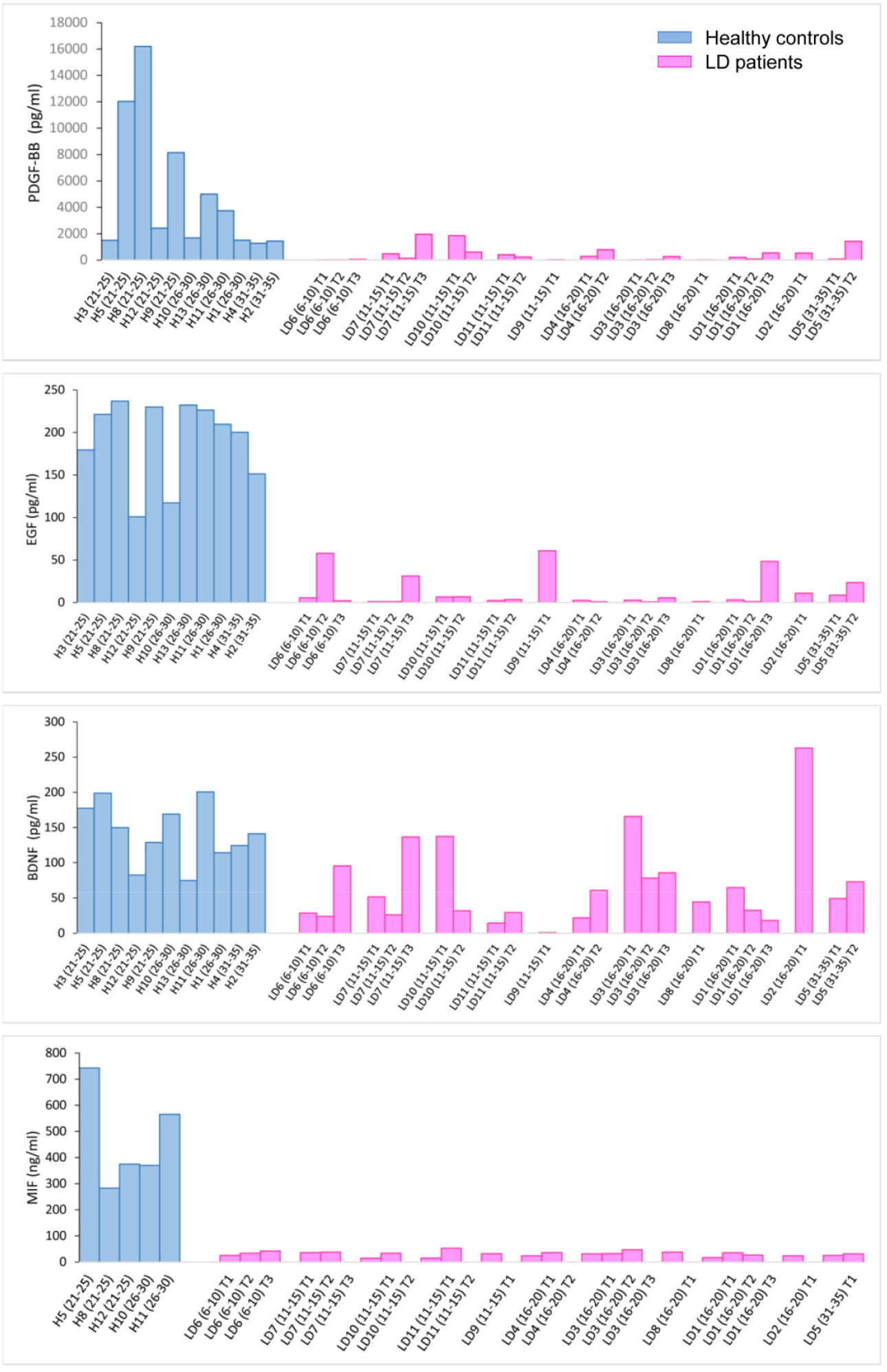
Longitudinal analysis of the levels of the PDGF-BB, EGF, BDNF, and MIF over time. (A-D) Plasma samples from patients collected at different times of exploration were evaluated using ELISA analysis and compared to the values found in samples from healthy controls. The number of the LD sample, the age range at the time of the extraction, and the number of the extraction are indicated in each case. Some patients provided three samples, while others contributed with only two or one. Results are expressed as absolute values and plotted according to age.

These results pointed out a better performance of PDGF-BB, EGF, and MIF as possible biomarkers of LD.

These analyses also demonstrated that in none of the cases, PDGF-BB **(Fig. 4A)**, EGF **(Fig. 4B)**, BDNF **(Fig. 4C)**, and MIF **(Fig. 4D)**, we found any correlation between the levels of these mediators and the progression of the disease in each patient (samples at latter stages of disease did not show consistent changes respect to values obtained in previous stages). In addition, values obtained at the first visit to the Neurology Department were mostly similar in all the patients, independently of the age of the patient, with some exceptions in the case of BDNF levels (**Supplementary Table S1**).

The results shown in **Fig. 4** also indicated that in the case of PDGF-BB, EGF, and MIF, there was no correlation between the levels of these mediators and the severity of the clinical presentation of the disease (**Supplementary Table S1 and S3)**. Patients with a severe presentation (e.g., LD-02 and LD-03 patients, in stage IV, according to the Delgado-Escueta scale (Kim et al., 2021)), showed lower levels of these mediators than controls, and the same was true in the case of patients with asymptomatic or with very weak presentation (e.g., LD-06 and LD-07 patients, in stage 0 of the Delgado-Escueta scale (Kim et al., 2021)). Moreover, no differences in clinical presentations and values of the analyzed mediators were found between patients carrying mutations in the *EPM2A* or *EPM2B* genes, or between males and females. Importantly, we found lower levels of PDGF-BB, EGF, and MIF in samples from LD patients that were asymptomatic or presented mild symptoms (e.g., LD-06, LD-07, in stage 0; **Supplementary Table S1 and S3**), suggesting the potential utility of these biomarkers for early diagnosis.

### 5. Specificity of PDGF-BB, EGF, BDNF, and MIF as biomarkers for LD

To determine whether the decreased levels of PDGF-BB, EGF, BDNF, and MIF were specific to LD, we analyzed samples from patients suffering from other monogenic epilepsies, including Dravet syndrome disease (caused by a pathogenic genetic variant in the sodium channel *SCN1A* gene) and GLUT1 deficiency syndrome (caused by a pathogenic genetic variant in the glucose transporter *SLC2A1* gene) (**Table 1**). As shown in **Fig. 5**, the three Dravet samples (OME-01 to OME-03) exhibited reduced levels of PDGF-BB (median of 1,067 pg/ml vs 2,424 pg/ml in healthy samples; **Fig. 5A**), EGF (median of 32 pg/ml vs 210 pg/ml in healthy samples; **Fig. 5B**), BDNF (median of 29 ng/ml vs 141 ng/ml in healthy samples; **Fig. 5C**) and MIF (47 ng/ml vs 375 ng/ml in healthy samples; **Fig. 5D**) (**Supplementary Table S1 and S3**). In the case of the two GLUT1 samples (OME-04 and OME-05), we found similar levels of PDGF in healthy samples (median 2,887 pg/ml vs 2,424 pg/ml in healthy samples), but lower levels of EGF (median 130 pg/ml vs 210 pg/ml in healthy samples), BDNF (median of 48 ng/ml vs 141 ng/ml in healthy samples) and MIF (median 121 ng/ml vs 375 ng/ml in healthy samples) in comparison with healthy samples **(Fig. 5A-D)** (**Supplementary Table S1 and S3**). Unfortunately, the reduced number of samples precluded us from regular statistical analysis of these models.

**Fig. 5.**
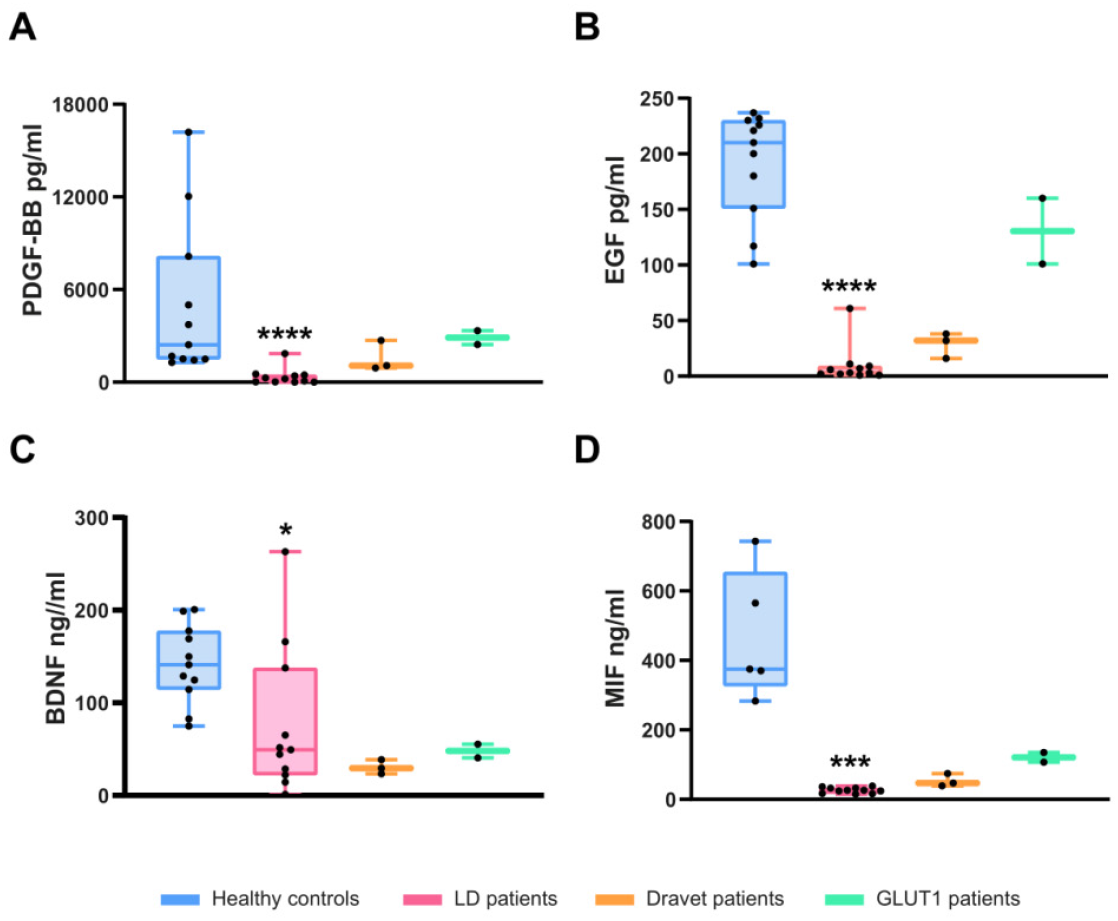
Comparison with other monogenic epilepsies. Plasma levels of PDGF-BB (A), EGF (B), BDNF (C), and MIF (D) were analyzed in three Dravet syndrome (pale orange) and two GLUT1 deficiency syndrome patients (pale green). For comparison, values from healthy controls and LD samples (as shown in Fig. 1) were included in the plot. Due to the limited sample size, statistical analysis to assess differences between genotypes could not be performed.

## DISCUSSION

One of the main challenges in the field of rare diseases is their delayed diagnosis. The identification of specific biomarkers has become an urgent need since they could facilitate both the diagnosis and prognosis of such conditions. The main goal of this study was to identify potential blood biomarkers of Lafora disease (LD). Using LD animal models, our group previously identified elevated levels of several chemokines and cytokines in the serum of *Epm2b-/-* mice (Rubio et al., 2023). However, when we analyzed the levels of these mediators (CXCL10, CCL20, and S100B) in plasma samples from a cohort of eleven LD patients, we found no statistical differences in comparison to the healthy controls. Probably differences in the phenotype presentation of the disease in mice and humans may account for this discrepancy. To expand our search for potential LD biomarkers, we conducted a high-throughput analysis using an array of 111 spotted antibodies targeting various cytokines, chemokines, and growth factors. This study identified eleven mediators whose levels were statistically different from LD patients and healthy controls. Interestingly, the most prominent differences, accompanied by robust statistical significance, corresponded to growth factors, including PDGF-AA/BB, EGF, BDNF, and MIF. In all these cases, we found a decrease in the levels of these mediators in LD samples in comparison to healthy controls. Notably, the levels of none of these markers were decreased in samples from *Epm2b-/-* mice (Rubio et al., 2023), pointing out again to differences between mice and human.

Then, we selected four of these markers, PDGF-AB/BB, EGF, BDNF, and MIF, and validated their levels by ELISA. Platelet-derived growth factor (PDGF) is a neurotrophic factor (NTF) produced by pericytes and endothelial cells (Cabezas et al., 2019). Several isoforms of this protein exist, including PDGFA and PDGFB, which form homo and heterodimers (PDGF-AA, PDGF-AB, and PDGF-BB) (Cabezas et al., 2019), (Sil et al., 2018). These ligands act on PDGF receptors (PDGFRs) expressed in endothelial cells, neurons, astrocytes, and microglia (Klement et al., 2019). As NTFs, they regulate neuronal function, and decreased PDGF-BB levels have been associated with neuronal loss and other disease-related outcomes (Bondarenko and Saarma, 2021). For instance, a PDGF-BB knockdown impairs hippocampal neurogenesis and increases susceptibility to chronic stress in mice (Li et al., 2023). Consequently, PDGF-BB is considered a peripheral marker of neurodegeneration (Hammerschmidt et al., 2023). Lower plasma PDGF-BB levels correlate with mild cognitive impairment in Alzheimer’s disease (AD), being considered PDGF-BB as one of the most important biomarkers associated with AD (Sil et al., 2018). Similarly, reduced PDGF-BB levels in plasma have been observed in severe ischemic stroke patients (Hutanu et al., 2020). In Pompe disease, a particular type of glycogen storage disorder due to impairments in the activity of the lysosomal alpha-glycosidase enzyme, lower levels of PDGF-BB in plasma are also detected, and they may be used to differentiate between asymptomatic and symptomatic patients (Fernandez-Simon et al., 2019). Notably, this latter case is particularly relevant to LD, as both conditions involve glycogen accumulation, although in LD polyglucosans accumulate in the cytosol rather than in the lysosome. Therefore, reduced levels of PDGF-BB in LD patients could indicate impaired neuronal functionality, neurodegeneration, and heightened hyperexcitability, as previously reported in animal models of LD (Rubio et al., 2023). Furthermore, PDGF-BB plays critical roles in angiogenesis, vessel stabilization, blood flow regulation, tissue repair, and blood-brain barrier (BBB) integrity. Deficient PDGF-BB levels disrupts these processes (Arango-Lievano et al., 2018), potentially contributing to the increased T-lymphocyte infiltration into the brain parenchyma that occurs in LD mice (Rubio et al., 2023). Since exogenous administration of PDGF-BB enhances vascular health and reduces spontaneous epileptiform activity (Arango-Lievano et al., 2018), we hypothesize that PDGF-BB treatment could benefit LD patients.

Epidermal growth factor (EGF) is another NTF involved in cell survival, proliferation, migration, and differentiation. It is synthesized in both the brain and peripheral tissues (Scalabrino, 2021), with its peripheral production gaining access to the central nervous system (CNS) through the blood-brain barrier, so the effect of EGF on the CNS is the sum of the CNS and peripheral production, the latter gaining access to CNS through the blood-brain barrier (BBB). EGF signals through the EGF receptor (EGFR), which is expressed on neurons, astrocytes, oligodendrocytes, and microglia, supporting neural stem cell maintenance, oligodendrocyte differentiation, and neuronal homeostasis (Scalabrino, 2022). In astrocytes, EGF regulates the glutamate-glutamine cycle by inducing glutamine synthase, which converts glutamate into glutamine for neuronal conversion to glutamate or GABA (Scalabrino, 2022). Reduced levels of EGF have been linked to demyelination, which could cause multiple sclerosis (MS) (Rivera et al., 2022). Decreased levels of EGF in plasma have also been reported in Parkinson’s (PD) and AD, limiting in this way EGFR activation (Rivera et al., 2022). Interestingly, EGF shares some signaling mediators with the PDGF pathway (Ben Jemii et al., 2020), and physical interactions between the EGFR and PDGFR signaling pathways have been proposed (Ben Jemii et al., 2020). These results are in agreement with our STRING analysis, which suggests a functional link between PDGF and EGF (**Fig. 1B**). Therefore, reduced levels of EGF in LD may contribute to neuronal dysfunction, neurodegeneration, and hyperexcitability.

Brain-derived neurotrophic factor (BDNF) is essential for neuronal plasticity, proliferation, differentiation, and connectivity. It functions also as a stress-responsive protection factor involved in learning and memory. BDNF signaling through the TrkB receptor promotes the expression of antioxidant enzymes, such as superoxide dismutase 2 (SOD2) and glutathione reductase, thereby protecting neurons and glial cells from excitotoxicity and oxidative damage (Cabezas et al., 2019). Decreased levels of BDNF in plasma have been reported in Huntington’s disease (HD) and Gaucher disease (GD) (Hammerschmidt et al., 2023). The reduced levels of BDNF observed in LD patients may reflect their heightened susceptibility to oxidative stress, as reported in LD mouse models (Roma-Mateo et al., 2015).

Finally, macrophage migration inhibitor factor (MIF) is primarily produced by T-lymphocytes but is also synthesized by endothelial cells and neurons. It has a dual role in cellular pathophysiology. On the one hand, MIF is considered a pro-inflammatory cytokine that recruits multiple inflammatory mediators, leading to the activation of microglia and astrocyte-derived neuroinflammation (Zeng et al., 2024). But, on the other hand, MIF has a neuroprotective role in defending neurons from oxidative stress and apoptosis (Zeng et al., 2024), (Xuan et al., 2023): in PD, MIF has a neuroprotective effect by suppressing inflammatory responses, inhibiting apoptosis, and inducing autophagy (Li et al., 2019), (Sumaiya et al., 2022), (Matejuk et al., 2024). Similarly, MIF protects against protein misfolding in stroke and amyotrophic lateral sclerosis (ALS) (Matejuk et al., 2024). Thus, the reduced levels of MIF in LD could exacerbate neurodegeneration and neuronal hyperexcitability.

Our longitudinal study reveals a significant reduction in these key neuroprotective growth factors (PDGF-BB, EGF, BDNF, and MIF) in the plasma of LD patients compared to healthy controls. Since in the case of BDNF, some values of LD samples were as high as some found in healthy controls, in our opinion, the levels of PDGF-BB, EGF, and MIF show a better performance as possible biomarkers of LD. Interestingly, reduced levels of these biomarkers were observed in asymptomatic patients, highlighting their potential utility as early diagnostic indicators.

In this work, we have also analyzed three samples from Dravet and two from GLUT1 deficiency patients. Although sample sizes were insufficient for robust statistical analysis, these samples also showed reduced levels of some of the markers found in LD patients, suggesting common pathophysiological mechanisms. Given the crucial roles that these proteins have in neuronal survival, function, and homeostasis, their depletion may directly contribute to the neuronal dysfunction, neurodegeneration, and hyperexcitability characteristic of these types of genetic epilepsies (LD, Dravet, and GLUT1 deficiency).

We are aware that the main limitation of this study is the reduced number of independent samples. This is a general limitation for rare diseases, especially in the case of ultra-rare diseases such as Lafora disease, with a prevalence of less than 4 patients per 1,000,000 individuals, where access to LD samples is quite complicated.

## Data Availability

All data produced in the present study are available upon reasonable request to the authors

## DECLARATIONS

### Data availability statement

The data that support the findings of this study are available from the corresponding author upon reasonable request.

## Acknowledgments and Funding statement

We want to acknowledge the donors and the CIBERER Biobank for their collaboration. We also thank Dr. Maria Adelaida García Gimeno for the critical reading of the manuscript and Ángela Campos-Rodríguez for the samples storage organization. This work was supported by grants from the Spanish Ministry of Science and Innovation (PID2023-148103OB-I00), la Fundació La Marató TV3 (202032), CIBERER (ACCI2020 and ACCI2023-03-742), and the Prometeo program from Generalitat Valenciana (CIPROM2022/42) to P.S.

## Conflict of interest disclosure

None of the authors have any conflict of interest to disclose.

## Ethics approval statement

The study design and sample collection were approved by the Ethics and Scientific Committees from Biobank-Centro de Investigación Biomédica en Red de Enfermedades Raras (CIBERER, Valencia, Spain) (Approval CEI-SP 20230707/05), from Fundación Jimenez Diaz (Madrid, Spain) (Approval LAF/BM/S-2019), from Hospital Vall d’Hebron (Barcelona, Spain) (Approval PR(AG)255/2023) and from Consejo Superior de Investigaciones Cientificas (CSIC, Madrid, Spain) (Approval 153/2024). Patient informed consent was obtained from all the individuals involved in this study.

## Author’s contribution

MME and MA performed the experiments. MME analyzed the data and participated in the writing of the manuscript. MM, LG, MS, JMS, LA, MQ, and MT participated in the collection of samples and assessed LD clinical presentations. PS analyzed the data and wrote the manuscript. All authors have read and approved the final version of the manuscript.

## Notes

### Competing Interest Statement

The authors have declared no competing interest.

### Funding Statement

This study was funded by grants from the Spanish Ministry of Science and Innovation (PID2023-148103OB-I00), la Fundacio La Marato TV3 (202032), CIBERER (ACCI2020 and ACCI2023-03-742), and the Prometeo program from Generalitat Valenciana (CIPROM2022/42) to P.S.

### Author Declarations

The study design and sample collection were approved by the Ethics and Scientific Committees from Biobank-Centro de Investigacion Biomedica en Red de Enfermedades Raras (CIBERER, Valencia, Spain) (Approval CEI-SP 20230707/05), from Fundacion Jimenez Diaz (Madrid, Spain) (Approval LAF/BM/S-2019), from Hospital Vall dHebron (Barcelona, Spain) (Approval PR(AG)255/2023) and from Consejo Superior de Investigaciones Cientificas (CSIC, Madrid, Spain) (Approval 153/2024). Patient informed consent was obtained from all the individuals involved in this study

