## Supplementary Material for "Identification of Plasma Growth Factors and Cytokines as Diagnostic Biomarkers for Lafora Disease"

**Supplementary Fig S1: Cytokine levels elevated in mouse models of LD are not differentially present in the plasma of LD patients in comparison to healthy controls.** Plasma levels of CXCL10 (A), S100B (B) and CCL20 (C) were analyzed in four samples from healthy subjects (pale blue) and seven samples from LD patients (pale pink) by using ELISA assays. Results are expressed as median with a range. Statistical differences between the groups were assessed using the Mann-Whitney non-parametric t-test. P-values are indicated.

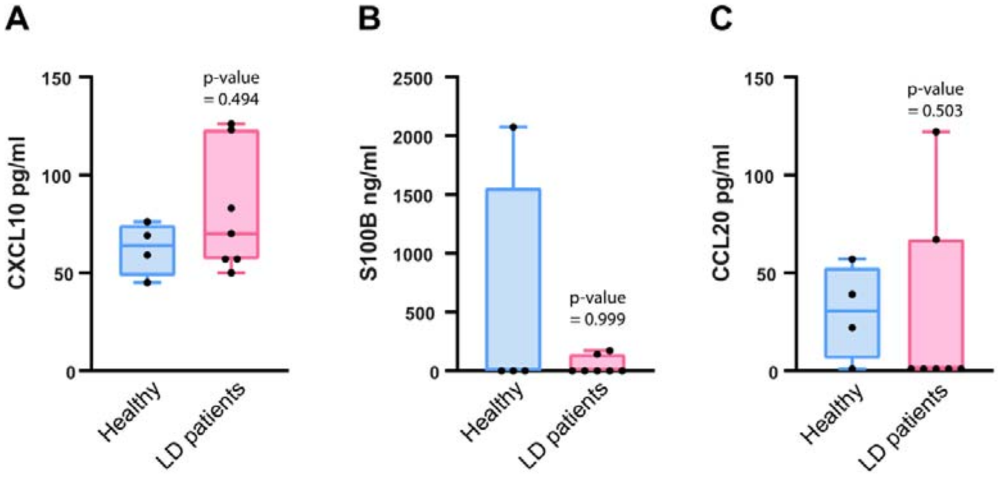

**Supplementary Table S1**. Description of the LD patients used in this work. The number of the patient, the affected gene and the mutation (both at DNA and protein level), gender, age at clinical onset and clinical presentation at different times of recruitment. The phenotytic stage of the patient according to Kim et al., 2021 is also indicated.

| Lafora Patient ID | Gene | Mutation | | SEX (M/F) | Age range at clinical onset (years) | First symptom | SEIZURES | | | Other seizures types at onset | Cognitive impairment at onset | Dementia at onset | Gait impairment at onset | Speech impairment at onset | T1 | | | | | | T2 | | | | | | T3 | | | | | |
|---|---|---|---|---|---|---|---|---|---|---|---|---|---|---|---|---|---|---|---|---|---|---|---|---|---|---|---|---|---|---|---|---|
| | | DNA | Protein | | | | GTC seizures at onset | Myoclonic seizures at onset | Visual seizures at onset | | | | | | AGE range T1 | CURRENT TREATMENT | GTCS FREQUENCY | MYOCLONIC SEIZURES FREQUENCY | OTHER SEIZURES FREQUENCY | Stage | AGE T2 | CURRENT TREATMENT | GTCS FREQUENCY | MYOCLONIC SEIZURES FREQUENCY | OTHER SEIZURES FREQUENCY | Stage | AGE T3 | CURRENT TREATMENT | GTCS FREQUENCY | MYOCLONIC SEIZURES FREQUENCY | OTHER SEIZURES FREQUENCY | Stage |
| **LD1** | EPM2A | c.877C>T homozygous | p.Gln293Ter | F | 6-10 | Visual seizures | Yes | Yes | Yes | Atonic, Absences | Yes | Yes | Yes | Yes | 16-20 | VPA, CZP, PER, metformin | 0 | 1/month | Visual: 1-3/month | **III** | 16-20 | VPA, CZP, PER, metformin | 0 | 1/week-1/month | Visual: 1-2/month; Atonic: 1/2-3 months | **III** | 16-20 | VPA, CZP, PER, metformin | 0 | Daily, uncountable | Atonic: 1/week-1/month; Absences: 1/week-1/month | **IV** |
| **LD2** | EPM2A | c.508C>T homozygous | p.Pro170Ser | M | 6-10 | Visual seizures | Yes | Yes | Yes | no | Yes | Yes | Yes | Yes | 16-20 | VPA, PER, BRV, atkins diet | 1/week | Daily, uncountable | Visual: 1/2-3 months | **IV** | sample not obtained | | | | | | sample not obtained | | | | | |
| **LD3** | EPM2B | c.205C>G homozygous | p.Pro69Ala | F | 11-15 | Visual and myoclonic seizures | Yes | Yes | Yes | absences, atonic | (diagnosed with ADHD) | Yes | Yes | Yes | 16-20 | VPA, LEV, ZNS, CZP | 1/month | Daily, uncountable | Visual: 1-2/month; Absences: daily and uncountable | **IV** | 16-20 | VPA, LEV, ZNS, CZP | 1-7/month | Daily, uncountable | Visual: 0-3/week; Absences: daily and uncountable; Atonic: 1/month | **IV** | 16-20 | VPA, LEV, ZNS, CZP, ketogenic diet | 5-11/month | Daily, uncountable | visual: 1/month; absences: daily and uncountable; Atonic: 5/6 months | **V** |
| **LD4** | EPM2A | c.721 C>T and exon 2 deletion | p.Arg241* | M | 11-15 | Cognitive | Yes | Yes | Yes | absences | Yes | no | no | no | 16-20 | VPA | 0 | 1/2-3 month | Visual: 1/month | **II** | 16-20 | VPA, CBD, TPM, metformin | 1/year | 1/month | Visual: 1/month | **II** | sample not obtained | | | | | |
| **LD5** | EPM2A | c.163C>T / c.835G>A | p.Gln55X / p.Gly279Ser | M | 21-25 | GTCS | Yes | Yes | Yes | no | no | no | no | no | 31-35 | VPA, CZP, metformin | 0 | 0 | no | **I** | 31-35 | VPA,CZP, metformin | 0 | 0 | no | **I** | sample not obtained | | | | | |
| **LD6** | EPM2A | c.259A>G homozygous | p.Lys87Glu | F | asymptomatic | no | no | no | no | no | no | no | no | no | 6-10 | no | no | no | no | **0** | 6-10 | no | no | no | no | **0** | 6-10 | no | no | no | no | **0** |
| **LD7** | EPM2A | c.259A>G homozygous | p.Lys87Glu | M | 1-5 | Cognitive | no | no | no | no | Yes | no | no | Yes | 11-15 | no | no | no | no | **0** | 11-15 | no | no | no | no | **0** | 11-15 | no | no | no | no | **0** |
| **LD8** | EPM2B | c.348C>A homozygous | p.Cys116Ter | F | 11-15 | GTC and visual seizures | Yes | Yes | Yes | no | Yes | Yes | Yes | Yes | 16-20 | VPA, LEV, CZP, ZNS, metformin | 1-4/month | Daily, uncountable | no | **IV** | sample not obtained | | | | | | sample not obtained | | | | | |
| **LD9** | EPM2A | c.834C>A / c.108_139del | p.Cys278* / p.Ala37fs | M | 11-15 | GTC and visual seizures | Yes | Yes | Yes | absences | Yes | Yes | Yes | Yes | 11-15 | ZNS, VPA, CLB, metformin | 0-5/month | Daily, uncountable | no | **II** | sample not obtained | | | | | | sample not obtained | | | | | |
| **LD10** | EPM2B | c.205C>G / c.1133T>C | p.Pro69Ala / p.Leu378Pro | F | 11-15 | GTCS | Yes | Yes | no | absences | no | no | no | no | 11-15 | VPA, metformin | 1/ year (she has had only 1 GTCS since clinical onset at that visit) | 1/day-1/week | absences: 1/week | **I** | 11-15 | VPA, CZP, metformin | 1/week - 1/month | Daily | no | **I** | sample not obtained | | | | | |
| **LD11** | EPM2B | c.205C>G / c.1133T>C | p.Pro69Ala / p.Leu378Pro | F | 11-15 | Myoclonic seizures | Yes | Yes | no | absences | Yes | no | no | no | 11-15 | VPA, CZP, metformin | 1/ year (she has had only 1 GTCS since clinical onset at that visit) | Daily | no | **I** | 11-15 | VPA, CZP, BRV, metformin | 1/week - 1/month | Daily, uncountable | no | **II** | sample not obtained | | | | | |

**M: male; F: female**
**BRV: brivaracetam; CBD: cannabidiol; CLB: clobazam; CZP: clonazepam; LEV: levetiracetam; PER: perampanel; TPM: topiramate; VPA: valproic acid; ZNS: zonisamid**

**GTCS: generalized tonic clonic seizures**

**ADHD: attention deficit hyperactivity disorder**

**Supplementary Table S2.** Plasma-EDTA samples from LD patients and healthy controls were analyzed using the Proteome Profiler Human Cytokine Array kit (R&D systems). This array consists of 111 different captured antibodies spotted on a nitrocellulose membrane for the detection of multiple cytokines, chemokines, growth factors, and other soluble proteins. The intensity of the signals was analyzed using the Quick Spot software provided by the manufacturer. The analysis of the differential expression between healthy and LD samples was performed by using the FLASKI software (http://flaski.age.mpg.de). In this work, we focus our attention on mediators with a fold change between healthy and LD samples greater than 2 or less than 0.5 for further analysis. Average intensity of the spots determined by the Quick Spot software in the Proteome Profiler Human Cytokine Array Kits. Eight healthy and eleven samples from independent LD patients were analyzed as described in Materials and Methods. Fold change of the Healthy vs LD samples and the corresponding p-values, calculated by using an unpaired non-parametric t-test (Mann-Whitney test) is also indicated. In the Table we highlight in orange the mediators with a fold change <0.5 or >2, with a p-value lower than 0.05. Mediators are ordered according to fold change. Values corresponding to Cxcl10 and Ccl20 are highligted in grey.

| **Healthy vs Patients** | **Average Healthy (n=8)** | **Patients (n=11)** | **Healthy vs LD Fold Change<0.5 or >2** | **p-value** |
|---|---|---|---|---|
| NC (Negative control) | 0,00 | 0,00 | | |
| PDGF-AB/BB | 9000,41 | 1065,69 | 8,45 | 0,0001 |
| PDGF-AA | 7197,94 | 1420,41 | 5,07 | 0,0009 |
| EGF | 2406,53 | 480,82 | 5,01 | 0,0008 |
| ENA-78/CXCL5 | 2292,20 | 720,45 | 3,18 | 0,0218 |
| TARC/CCL17 | 1128,13 | 399,52 | 2,82 | 0,0180 |
| BDNF | 1792,01 | 642,62 | 2,79 | 0,0011 |
| MIF | 4376,88 | 1787,67 | 2,45 | 0,0002 |
| Thrombospondin-1 | 5575,32 | 2360,47 | 2,36 | 0,0090 |
| Angiopoietin-1 | 1850,48 | 821,64 | 2,25 | 0,0146 |
| RANTES/CCL5 | 14307,91 | 6510,20 | 2,20 | 0,0707 |
| Dkk-1/Dickkopf-1 | 1268,62 | 584,47 | 2,17 | 0,0157 |
| Growth Hormone | 2559,68 | 1238,98 | 2,07 | 0,4164 |
| Serpin E1 | 15889,21 | 8103,80 | 2,00 | 0,0063 |
| CD31 | 18348,39 | 12031,88 | 1,52 | 0,1199 |
| IL-2 | 391,90 | 267,47 | 1,47 | 0,2843 |
| IL-10 | 357,41 | 256,53 | 1,39 | 0,3196 |
| IL-3 | 323,89 | 232,67 | 1,39 | 0,3555 |
| TGF-alpha | 264,78 | 191,22 | 1,38 | 0,4326 |
| EMMPRIN | 8300,32 | 6101,65 | 1,36 | 0,1364 |
| VEGF | 420,22 | 314,19 | 1,34 | 0,2412 |
| I-TAC | 275,11 | 217,39 | 1,27 | 0,3339 |
| PF4 | 21262,51 | 16987,93 | 1,25 | 0,2545 |
| MCP-1 | 698,33 | 558,68 | 1,25 | 0,2874 |
| Fas Ligand | 686,93 | 554,64 | 1,24 | 0,5538 |
| IL-1beta | 111,63 | 91,02 | 1,23 | 0,7483 |
| GM-CSF | 436,19 | 360,91 | 1,21 | 0,3430 |
| Vitamin D BP | 10864,33 | 9102,84 | 1,19 | 0,4316 |
| Lipocalin-2 | 9882,83 | 8477,77 | 1,17 | 0,4670 |
| FGF basic | 992,51 | 853,65 | 1,16 | 0,4216 |
| IL-11 | 688,08 | 598,57 | 1,15 | 0,4238 |
| FGF-7 | 174,98 | 153,65 | 1,14 | 0,7732 |
| Cystatin C | 6093,82 | 5390,25 | 1,13 | 0,5752 |

| | | | | |
|---|---|---|---|---|
| MCP-3 | 154,34 | 139,17 | 1,11 | 0,8269 |
| TFF3 | 7729,50 | 6988,91 | 1,11 | 0,5960 |
| IL-13 | 66,18 | 59,91 | 1,10 | 0,8755 |
| IL-22 | 516,29 | 468,56 | 1,10 | 0,6680 |
| IL-27 | 281,25 | 256,19 | 1,10 | 0,8060 |
| Cripto-1 | 299,85 | 277,41 | 1,08 | 0,7528 |
| IL-32 | 219,52 | 204,49 | 1,07 | 0,8368 |
| Flt-3 Ligand | 244,06 | 227,41 | 1,07 | 0,7906 |
| GRO-alpha | 238,85 | 222,56 | 1,07 | 0,8229 |
| MMP-9 | 15859,98 | 14827,37 | 1,07 | 0,7042 |
| TNF-alpha | 652,80 | 616,97 | 1,06 | 0,8527 |
| G-CSF | 195,18 | 186,68 | 1,05 | 0,9013 |
| Angiopoietin-2 | 902,95 | 864,41 | 1,04 | 0,8443 |
| CD40 ligand | 2434,73 | 2330,87 | 1,04 | 0,8003 |
| SHBG | 10524,95 | 10123,08 | 1,04 | 0,8137 |
| IL-17A | 2350,17 | 2261,80 | 1,04 | 0,8087 |
| IL-4 | 462,03 | 452,13 | 1,02 | 0,9034 |
| TfR | 3972,84 | 3948,10 | 1,01 | 0,9460 |
| M-CSF | 308,06 | 307,14 | 1,00 | 0,9880 |
| IL-6 | 398,52 | 399,57 | 1,00 | 0,9902 |
| C-Reactive Protein | 24060,17 | 24255,02 | 0,99 | 0,9523 |
| IGFBP-2 | 7508,16 | 7700,11 | 0,98 | 0,8607 |
| RAGE | 755,71 | 777,83 | 0,97 | 0,9135 |
| IL-34 | 80,09 | 82,80 | 0,97 | 0,9415 |
| HGF | 332,92 | 344,46 | 0,97 | 0,8954 |
| IL-19 | 389,85 | 405,23 | 0,96 | 0,8538 |
| Complement Component C5/C5a | 7221,63 | 7523,88 | 0,96 | 0,7021 |
| IL-1ra | 253,18 | 267,50 | 0,95 | 0,8693 |
| SDF-1alpha | 2842,56 | 3006,60 | 0,95 | 0,6945 |
| Reference Spots | 19042,48 | 20161,79 | 0,94 | 0,2931 |
| FGF-19 | 1843,56 | 1968,06 | 0,94 | 0,7462 |
| ST2 | 2140,07 | 2295,75 | 0,93 | 0,7729 |
| Kallikrein 3 | 1082,75 | 1185,68 | 0,91 | 0,6796 |
| IL-31 | 129,15 | 141,44 | 0,91 | 0,8449 |
| RBP-4 | 25704,58 | 28187,33 | 0,91 | 0,2505 |
| Chitinase 3-like 1 | 8507,89 | 9348,44 | 0,91 | 0,5968 |
| Myeloperoxidase | 514,78 | 565,93 | 0,91 | 0,6994 |
| MIG | 189,08 | 209,09 | 0,90 | 0,7377 |
| MIP-1alpha/MIP-1beta | 103,79 | 114,90 | 0,90 | 0,8182 |
| IFN-gamma | 496,76 | 550,25 | 0,90 | 0,7270 |
| IGFBP-3 | 10917,62 | 12109,98 | 0,90 | 0,2035 |
| IP-10/Cxcl10 | 500,24 | 557,10 | 0,90 | 0,8025 |
| Apolipoprotein A-I | 13499,03 | 15043,87 | 0,90 | 0,1510 |
| Osteopontin | 10506,06 | 11736,96 | 0,90 | 0,5789 |
| Adiponectin | 31728,76 | 35476,32 | 0,89 | 0,3049 |
| VCAM-1 | 15482,02 | 17517,03 | 0,88 | 0,1261 |
| ICAM-1 | 2569,58 | 2907,73 | 0,88 | 0,3953 |
| Reference Spots | 17661,36 | 20034,80 | 0,88 | 0,0936 |
| Reference Spots | 17588,28 | 20031,55 | 0,88 | 0,1631 |
| IL-1alpha | 556,35 | 633,69 | 0,88 | 0,5997 |
| IL-12 p70 | 222,71 | 255,48 | 0,87 | 0,6310 |

| | | | | |
|---|---|---|---|---|
| IL-16 | 140,62 | 161,38 | 0,87 | 0,6350 |
| IL-24 | 152,79 | 176,11 | 0,87 | 0,7165 |
| Angiogenin | 25792,29 | 29859,86 | 0,86 | 0,1652 |
| Pentraxin-3 | 1360,08 | 1577,15 | 0,86 | 0,4438 |
| CD14 | 2957,66 | 3447,92 | 0,86 | 0,2352 |
| Resistin | 2242,92 | 2630,73 | 0,85 | 0,4640 |
| uPAR | 1039,76 | 1257,79 | 0,83 | 0,2304 |
| Complement Factor D | 5417,08 | 6555,63 | 0,83 | 0,1382 |
| Relaxin-2 | 324,35 | 395,80 | 0,82 | 0,2573 |
| IL-5 | 109,84 | 136,05 | 0,81 | 0,6881 |
| IL-18 BPa | 9468,18 | 11763,38 | 0,80 | 0,0453 |
| IL-15 | 52,10 | 64,95 | 0,80 | 0,6311 |
| IL-23 | 212,67 | 270,58 | 0,79 | 0,4648 |
| CD30 | 532,04 | 684,94 | 0,78 | 0,2812 |
| MIP-3alpha/Ccl20 | 199,75 | 258,50 | 0,77 | 0,2040 |
| IL-33 | 56,02 | 72,84 | 0,77 | 0,5874 |
| Leptin (antes sí) | 6051,72 | 7941,01 | 0,76 | 0,6168 |
| LIF | 118,66 | 158,21 | 0,75 | 0,5503 |
| IL-8 | 402,26 | 538,16 | 0,75 | 0,2277 |
| Endoglin | 8078,73 | 10960,22 | 0,74 | 0,0557 |
| BAFF | 3210,84 | 4392,92 | 0,73 | 0,0970 |
| DPPIV | 12368,52 | 17106,04 | 0,72 | 0,0768 |
| TIM-3/HAVCR2 | 4317,44 | 6438,51 | 0,67 | 0,0330 |
| MIP-3beta | 332,53 | 523,50 | 0,64 | 0,1826 |
| GDF-15/MIC-1 | 1795,33 | 3690,24 | 0,49 | 0,0052 |

**Supplementary Table S3**. Values of EGF, PGDF-BB, BDNF, and MIF determined by ELISA assays in LD samples at different recruitment times (T1, T2 and T3), in healthy samples and in other monogenic epileptic patients. F, female; M, male; nd, not determined; ns, no sample obtained.

| Lafora Patients | Age range (T1/T2/T3) | Sex | Gene | T1/T2/T3 | | | |
|---|---|---|---|---|---|---|---|
| | | | | EGF (pg/ml) | PDGF-BB (pg/ml) | BDNF (ng/ml) | MIF (ng/ml) |
| **LD1** | 16-20 | F | EPM2A | 3/1/49 | 215/83/547 | 65/33/18 | 17/36/27 |
| **LD2** | 16-20 | M | EPM2A | 11/ns/ns | 531/ns/ns | 263/ns/ns | 24/ns/ns |
| **LD3** | 16-20 | F | EPM2B | 3/1/6 | 1/32/262 | 165/78/85 | 32/33/47 |
| **LD4** | 16-20 | M | EPM2A | 2/1/ns | 273/784/ns | 22/61/ns | 24/36/ns |
| **LD5** | 31-35 | M | EPM2A | 9/24/ns | 85/1424/ns | 49/73/ns | 26/32/ns |
| **LD6** | 6-10 | F | EPM2A | 6/58/2 | 8/1/64 | 28/24/95 | 26/34/43 |
| **LD7** | 11-15 | M | EPM2A | 1/1/31 | 476/140/1953 | 51/26/136 | 36/38/37 |
| **LD8** | 16-20 | F | EPM2B | 1/ns/ns | 15/ns/ns | 44/ns/ns | 38/ns/ns |
| **LD9** | 11-15 | M | EPM2A | 61/ns/ns | 29/ns/ns | 1/ns/ns | 32/ns/ns |
| **LD10** | 11-15 | F | EPM2B | 7/7/ns | 1848/600/ns | 137/31/ns | 15/34/ns |
| **LD11** | 11-15 | F | EPM2B | 2/3/ns | 411/229/ns | 14/29/ns | 16/53/ns |
| **Healthy Controls** | | | | | | | |
| **H1** | 26-30 | F | | 210 | 1500 | 114 | nd |
| **H2** | 31-35 | M | | 151 | 1433 | 141 | nd |
| **H3** | 21-25 | F | | 180 | 1488 | 178 | nd |
| **H4** | 31-35 | M | | 200 | 1277 | 125 | nd |
| **H5** | 21-25 | F | | 221 | 12037 | 199 | 743 |
| **H6** | 26-30 | M | | nd | nd | nd | nd |
| **H7** | 21-25 | F | | nd | nd | nd | nd |
| **H8** | 21-25 | M | | 237 | 16192 | 150 | 283 |
| **H9** | 21-25 | F | | 230 | 8151 | 129 | nd |
| **H10** | 26-30 | F | | 117 | 1681 | 170 | 370 |
| **H11** | 26-30 | M | | 226 | 3734 | 201 | 565 |
| **H12** | 21-25 | F | | 101 | 2424 | 83 | 375 |
| **H13** | 26-30 | M | | 232 | 5004 | 75 | nd |
| **Other monogenic epileptic patients** | | | | | | | |
| **OME1** | 16-20 | M | (SCNA1) | 16 | 915 | 23 | 47 |
| **OME2** | 16-20 | M | (SCNA1) | 32 | 1067 | 29 | 39 |
| **OME3** | 26-30 | M | (SCNA1) | 38 | 2702 | 39 | 74 |
| **OME4** | 26-30 | M | (SLC2A1) | 101 | 2439 | 41 | 107 |
| **OME5** | 26-30 | M | (SLC2A1) | 160 | 3334 | 55 | 135 |
| | | | **Median LD values (T1/T2/T3)** | **03/ 03/ 18** | **215/ 185/ 405** | **49/ 32/ 88** | **26/ 35/ 40** |
| | | | **Median control values** | **210** | **2424** | **141** | **375** |
| | | | **Median Dravet values** | **32** | **1067** | **29** | **47** |
| | | | **Median GLUT1 values** | **130** | **2887** | **48** | **121** |
